# IMPACT OF BK POLYOMAVIRUS INFECTION IN CHILDREN AFTER KIDNEY TRANSPLANT IN A REFERENCE CENTER IN SALVADOR – BAHIA, BRAZIL

**DOI:** 10.1101/2024.12.19.24319177

**Authors:** Mariana Perdiz Fucs Machado, Ricardo José Costa Mattoso

## Abstract

**Introduction:** BK Polyomavirus infection has a high incidence in the post-renal transplant phase and is considered a significant cause of premature graft failure after transplantation. Pediatric renal patients undergo relatively more potent immunosuppression; however, despite the large number of studies conducted in adult renal recipients, there is limited information on BKPyV infection in the pediatric population.

**Objective:** To estimate the incidence of BK Polyomavirus infection after kidney transplantation in pediatric patients undergoing immunosuppressive therapy at a referral center.

**Methodology:** Observational descriptive study, with data collection based on medical records of children undergoing kidney transplantation at Hospital Ana Nery, Salvador-BA, from 2009 to 2017.

**Results:** Forty-one children were evaluated, of which 21 (51.2%) were female. The mean age of the sample was 11 ± 3, with a higher frequency of patients in the age range of 13-17 years (43.9%). Regarding the presence of BKPyV infection, most patients were not infected (56.1%). Regarding the type of antiproliferative and calcineurin inhibitor, Mycophenolate Mofetil was used in 37 (90.2%) patients and Tacrolimus in 40 (97.6%). When comparing the groups with the presence or absence of BKPyV infection, there was no statistically significant difference between the variables: sex (p = 0.890), age (p = 0.829) and type of antiproliferative used (p = 0.187). In the analysis of renal function, an increase in creatinine was observed in 38.9% of infected patients, while among the uninfected patients, only 13% presented an increase in the period of one year. The difference between these two groups (p = 0.05) therefore demonstrated an association between worsening renal function and the presence of BKPyV infection.

**Conclusion:** The incidence of BK Polyomavirus infection in pediatric patients undergoing kidney transplantation, in the period of one year after the surgical procedure, is almost 50%. No differences could be demonstrated between the groups with and without BKPyV infection and established risk factors, such as sex, age and immunosuppressive therapy used. Furthermore, a worsening of renal function was evidenced among infected patients, when compared to the uninfected group, in this period of one year.

## 1. INTRODUCTION

BK polyomavirus (BKPyV) is a small DNA virus first described in 1971 after the discovery of infected cells in the urine of a renal transplant recipient with ureteral stricture. With the widespread use of potent immunosuppressive agents and improved viral surveillance practices, it has emerged as an important cause of graft loss in renal transplant recipients.1

BK polyomavirus replication occurs during immunosuppressive states, and BKPyV infection in immunocompromised individuals can lead to complications such as urethral stricture and BKV-associated nephropathy (BKVN), which is rare in non-renal transplant settings. The development of BKVN occurs in approximately 1% to 10% of cases and is associated with graft loss rates of approximately 50%. However, the introduction of screening programs allows earlier intervention and control of viremia before the onset of interstitial disease, reducing this value.1,2

Viremia rates among kidney transplant recipients vary widely in the literature, with prevalence between 1.5 and 33%, and peak incidence in the first year after surgery. Viruria, on the other hand, has a prevalence of approximately 25% in transplant patients. Regarding renal function, studies describe higher creatinine levels in viruric and viremic patients, and this renal dysfunction was assessed in a 24-month follow-up by the authors who reached these results.1,3

Despite the large number of studies conducted in adult kidney recipients, there is limited information on BKPyV infection in pediatric renal patients. Knowledge of the prevalence of BK polyomavirus in the pediatric population is important, given the relatively more potent immunosuppression used. Furthermore, children are more likely to receive a kidney from a BK-positive donor at transplantation, and thus younger seronegative children may be at risk of a devastating primary infection in this setting.4,5,9

Currently, no established antiviral treatment is available, and control of viral infection is tentatively achieved by reducing immunosuppression. Given that late diagnosis of BKVN is usually associated with irreversible decline in graft function and that most patients with viremia will eventually progress to BKVN, regular screening for BKV reactivation, particularly during the first two years after transplantation, with subsequent preventive reduction of immunosuppression, represents the procedure most commonly adopted by transplant centers. However, the risk of graft rejection due to reduced doses of immunosuppressive drugs suggests that preventive treatment should be applied only to selected groups of patients at high risk of developing BKV-related complications. Means of identifying these high-risk patients therefore need to be investigated.2,4

There is no clear evidence to support any specific modification of immunosuppressive therapy. In vitro analyses, however, suggest that reduction or withdrawal of calcineurin inhibitors should be the first step in modifying immunosuppression because of their effect on T lymphocytes. Reduction or withdrawal of antiproliferative drugs, particularly mycophenolate, is also a common target for changes in immunosuppressive regimen, since the presence of MMF in baseline immunosuppressive regimens has been described as a significant risk factor for BKV infection. Thus, despite the lack of controlled studies, it seems reasonable, at least for patients at lower risk of rejection, to withdraw or reduce the use of tacrolimus and mycophenolate by about 25 to 50%. After immunosuppression is reduced, renal function must be carefully monitored due to the risk of rejection.2,4

Thus, the central question of the research is to focus on the incidence of BK polyomavirus in the pediatric population and the consequences of this infection for the transplanted graft, with an assessment of the use of immunosuppressants and failure of the affected kidney. With this data, the multidisciplinary team at the health center can gather information about the needs and deficiencies of individuals in the post-transplant phase.

## 2. OBJECTIVES

### 2.1. General objective

To estimate the incidence of BK polyomavirus infection after kidney transplantation in pediatric patients undergoing immunosuppressive therapy.

### 2.2. Specific objectives

2.2.1. To estimate the frequency of patients infected with polyomavirus according to the immunosuppressant used.

2.2.2. To estimate the frequency of patients infected with polyomavirus who presented worsening of renal function.

2.2.3. To evaluate the impact on renal function over a one-year period.

## 3. BACKGROUND

### 3.1. Kidney transplant

Kidney transplant is a surgical procedure that offers the possibility of restoring the health and quality of life of chronic kidney patients. The main concern after transplantation is rejection. The cells of the immune system identify the transplanted organ as something foreign and try to destroy it. To avoid rejection by the body itself, it is necessary to use immunosuppressive medication, which will act on the immune system, preventing the recognition of the transplanted organ and its rejection.6

Kidney transplantation (KT) is the best form of treatment for end-stage renal disease in childhood, and current results on patient and graft survival indicate that this form of treatment should be encouraged, even in young children.6

The major difference between transplantation in adults and in children is related to the response of the child’s immune system, which, because it is in the process of maturation, requires more intense immunosuppression. Immunosuppression weakens the immune system in order to improve transplant acceptance. On the other hand, this weakening also makes the patient more susceptible to opportunistic infections. Given that the dose and potency of immunosuppressants are higher for pediatric transplant recipients, they are more likely to suffer from post-transplant infections.6

### 3.2. Characteristics of Polyomavirus and Infection Polyomaviruses belong to the Polyomaviridae family

Within this family, the JC (JCV) and BK (BKV) polyomaviruses stand out because they are capable of causing infections specifically in humans. JCV and BKV are small viruses, measuring approximately 45 nm in diameter, with circular, double-stranded, non-enveloped DNA.2

Polyomaviruses can cause latent infections that can last a lifetime, in both adults and children. In immunocompetent individuals, infection is asymptomatic and does not cause problems. However, in transplant patients undergoing immunosuppression, both a primary infection and reactivation of BKV can cause dysfunction, placing the newly transplanted renal graft at risk. BK infection is predominant (> 90%) in the first two years after transplantation, especially in the first trimester. Screening initiatives have focused mainly on the first 6-12 months.7

The mode of viral transmission responsible for primary infection is still unknown. Although BKV can rarely be recovered from the respiratory tract, the rapid acquisition of antibodies in the first years of life suggests that transmission occurs by this route. Recently, transplacental transmission has also been proposed. Other body fluids may also be potentially involved in the transmission of BKV infections, such as nasopharyngeal aspirates from children hospitalized with severe respiratory infections, as well as semen and blood from healthy donors. BKV DNA can also be isolated from tissues of the genital tract and normal skin. Since the viral genome has often been detected in healthy kidneys, organ donors may also be an important vehicle for viral transmission during transplantation.7

As for the process of reactivation of the infection, it is more restricted to moments of immunosuppression and malignant diseases, situations in which the immune system is deficient. BKV reactivates in the urinary tract and its first manifestation is the excretion of the virus in the urine, which is clinically asymptomatic. Depending on the intensity of viral replication, the virus may be eliminated in the urine, without invading the bloodstream or progressing to a viremic state and establishing the disease.6

In kidney transplantation, the prevalence of latent polyomavirus infection is high. BKV infection has been reported in up to 65% of kidney recipients. However, BKV disease usually occurs in approximately 10% of this patient population. Manifestations of this type of infection after kidney transplantation are usually: development of interstitial nephritis, ureteral stenosis, systemic infection or bladder cancer. Polyomavirus disease should be suspected when a kidney recipient presents with macroscopic hematuria, urinary tract obstruction or persistently increased creatinine levels without apparent cause.7

### 3.3. Use of immunosuppressants

The combined use of immunosuppressants is part of the treatment of pediatric patients undergoing kidney transplantation. In the period leading up to the transplant, induction therapy occurs, in which immunosuppressive medications are administered to reduce the risk of rejection even after the surgical procedure has been performed.8

The use of immunosuppressive medications makes graft recipients more susceptible to infections. In the case of viral infections, the most common are caused by cytomegalovirus, associated with herpes simplex, and polyomavirus BK.9

In the first weeks after transplantation, the use of immunosuppressants is associated with increased morbidity and mortality due to infectious complications. There is also the influence of inadequate use of immunosuppressant drugs by the patient, the effect of compatibility in the HLA (Human Leukocyte Antigen) system and its association with the risk of rejection of the kidney graft. After kidney transplantation, hospital infections associated with the surgical procedure itself as well as with the use of urinary devices may occur. Infections in the first month are generally not due to transmission by the graft, and are caused by viruses, bacteria and fungi originating from the hospital environment. From the beginning of the second month, viruses prevail over bacteria.10

The regimens used in immunosuppression include glucocorticoids, calcineurin inhibitors, antiproliferative agents, and monoclonal/polyclonal antibodies.8 Azathioprine is an antiproliferative immunosuppressant that inhibits humoral and cellular immunity and interferes with the differentiation of activated lymphocytes. It also reduces the production of macrophages and neutrophils and thus decreases the inflammatory activity of nonspecific components. The mechanism of action is the inhibition of enzymes that form adenosine monophosphate and guanosine, in addition to slowing down the biosynthetic pathway of purines. Azathioprine is effective in preventing kidney graft rejection, but is ineffective when the rejection process is already underway.8 14

Another antiproliferative agent used is mycophenolate sodium or mofetil, which inhibits purine synthesis, blocking the proliferation of activated T and B cells, i.e., decreasing the humoral and cellular response. Its use, despite reducing acute graft rejection, does not reduce chronic rejection rates. There are associations with increased rates of viral infections, but some can be prevented by prophylaxis.8

Among the factors that represent a greater risk for the development of BKN (BKV Interstitial Nephritis) is the intensity of immunosuppression, especially with the use of drugs such as mycophenolate mofetil and tacrolimus. The latter, a potent calcineurin inhibitor, used as prophylaxis and treatment of episodes of severe acute rejection, has been strongly associated with the development of polyomavirus disease, being used in the immunosuppressive regimen of up to 70% of cases of patients with BKN.7

### 3.4. Diagnosis of polyomavirus and renal graft loss

Polyomavirus type BK is the most common type of polyomavirus that affects renal grafts (Polyomavirushominis type I). It is found in the urine of 20% of healthy post-transplant patients, suggesting that it is a reactivation of an infection that occurred in childhood. Most infections are detected in the first year after transplantation, with tropism for Bowman’s capsule.

Diagnosis can be made by urinary cytology, renal biopsy and by PCR (polymerase chain reaction) of the viral load in urine and blood. In urinary cytology, infected cells are called decoy cells. In renal biopsy, pathological cells are seen in distal segments and in collecting ducts of the medulla. Renal dysfunction is caused by tubular necrosis and varying degrees of interstitial inflammation. The patterns that are associated with graft loss in histology are divided into three grades: A (viral cytopathic changes), B (viral cytopathic changes with varying degrees of inflammation, interstitial fibrosis, and tubular atrophy), and C (severe inflammatory changes and tubulointerstitial chronicity).11 The occurrence of dysfunction and acute rejection is associated with reduced graft and patient survival. Induction immunosuppression with the use of Thymoglobulin reduced the incidence of acute rejection to approximately 5%. However, this immunosuppression regimen increases the incidence rates of death due to infection. Renal graft survival rates after transplantation (6 months to 1 year) are 94.5% and 93.2% versus 97.6% and 97.5%, respectively, comparing patients with and without an episode of infection.12

Renal biopsy is the gold standard for identifying causes of renal graft lesions. The search for infectious antibodies, such as cytomegalovirus and polyomavirus, in addition to markers for lymphoproliferative disease of the transplant, can be performed by immunohistochemical study.12 The prognosis of BKN is directly related to the early diagnosis. Patients diagnosed at an early stage of infection and appropriately managed result in good recovery of graft function, however, patients diagnosed late end up losing the transplanted organ. The prevalence of graft loss in patients diagnosed with BKN ranges from 45% to 70%.7

## 4. METHODS

### 4.1. Study design

A descriptive observational study was conducted at the Ana Nery Hospital, Salvador - BA, with retrospective data collection based on information from medical records.

### 4.2. Study population

Data collection was performed at the hospital level in the nephrology department of the Ana Nery Hospital, Salvador-BA. The following inclusion criteria were defined: patients aged 5 to 17 years, of both sexes, who underwent kidney transplantation between 2009 and 2017. Cases in which there was missing information in the medical records to meet the causality criteria and cases of patients who presented reduced immunity not caused by immunosuppressive drugs were excluded.

### 4.3. Data source and operationalization of the collection

The physicians were responsible for analyzing the patients’ medical records and evaluating the data obtained in the research. The recruitment period was between July and November 2018. The basis for this research was the detection in the medical records of polyomavirus infection through PCR viruria tests and renal biopsy in children who had undergone transplantation between 1 month and 1 year after surgery. The results obtained were cross-referenced in a database to describe the characteristics of the incidence of BK polyomavirus infection in children after kidney transplantation and, consequently, these data were associated with sex, age, antiproliferative and calcineurin inhibitor used, worsening of renal function (with the criterion of creatinine elevation by 1.5 times the value prior to the diagnosis of the infection) and impact on renal function in the period of one year. 17 4.4. Variables: The following will be analyzed as quantitative variables: age (numerical), creatinine in the 1st month (mg/dL), creatinine in the 3rd month (mg/dL) and creatinine in the 12th month (mg/dL). In addition, the following will be evaluated as qualitative variables: BK polyomavirus infection (yes or no), sex (female or male), type of antiproliferative agent used (Azathioprine or Mycophenolate), calcineurin inhibitor used (Tacrolimus or Cyclosporine) and worsening of creatinine (yes or no). 4.5. Data analysis plan: After preparing the Database in the Excel Program, the categorical variables were expressed in absolute values and relative frequencies (percentages) and the quantitative variables in means and standard deviation according to the assumptions of normality, using the Kolmogorov-Smirnov test. The chi-square test was used to verify statistically significant differences in categorical variables, and the Student’s t-test was used for quantitative variables. Values of p < 0.005 were considered statistically significant. The analyses were conducted using the IBM Statistical Package for the Social Sciences (SPSS®, Chicago, IL, USA) V 14 software.

### 4.4. Variables

The following will be analyzed as quantitative variables: age (numerical), creatinine in the 1st month (mg/dL), creatinine in the 3rd month (mg/dL) and creatinine in the 12th month (mg/dL). In addition, the following will be evaluated as qualitative variables: BK polyomavirus infection (yes or no), sex (female or male), type of antiproliferative agent used (Azathioprine or Mycophenolate), calcineurin inhibitor used (Tacrolimus or Cyclosporine) and worsening of creatinine (yes or no). 4.5.

Data analysis plan: After preparing the Database in the Excel Program, the categorical variables were expressed in absolute values and relative frequencies (percentages) and the quantitative variables in means and standard deviation according to the assumptions of normality, using the Kolmogorov-Smirnov test. The Chi-Square test was used to verify statistically significant differences in categorical variables, and the Student’s T-test was used for quantitative variables. Values of p < 0.005 were considered statistically significant. The analyses were conducted using the IBM Statistical Package for the Social Sciences (SPSS®, Chicago, IL USA) V 14 software.

### 4.5. Data analysis plan

After preparing the database in Excel, the categorical variables were expressed in absolute values and relative frequencies (percentages) and the quantitative variables in means and standard deviations according to the assumptions of normality, using the Kolmogorov-Smirnov test. The chi-square test was used to verify statistically significant differences in the categorical variables and the Student’s t-test for the quantitative variables. Values of p < 0.005 were considered statistically significant. The analyses were conducted using the IBM Statistical Package for the Social Sciences (SPSS®, Chicago, IL USA) V 14 software.

### 4.6. Ethical aspects

This is a study in which risks are minimal because it is observational, with evaluation of medical records and analysis of existing data. Confidentiality regarding the identity of patients will be maintained, in addition to the preservation and confidentiality of the data studied. The benefits are related to the contribution of the results to better manage patients using immunosuppressive medications in the post-renal transplant phase. The research protocol was approved by the Ethics and Research Committee of the Ana Nery Hospital through Opinion no. 2.644.625 of 05/08/18 (Annex), in order to ensure the suitability of the project, as well as to guarantee the researchers’ commitment to preserving the anonymity of the patients involved in the work. The research was conducted in compliance with Resolution 466-12-CONEP-CNS-MS.

## 5. RESULTS

The sample of this study consisted of 41 pediatric patients (Appendix A) who underwent kidney transplantation, 51.2% of whom were female and 48.8% were male. The majority of patients in the study were in the age range of 13-17 years, corresponding to 43.9% of the total. The average age found was 11 +/- 3 (Table 1).

**Table 1.** Distribution of the sample of children undergoing kidney transplantation in a referral hospital according to sex and age. Salvador· BA, 2009 to 2017.

| Variables |  | n | % |
| --- | --- | --- | --- |
| Sex |  |  |  |
|  | Masculine | 20 | 48.8 |
|  | Feminine | 21 | 51.2 |
|  | **Total** | **41** | **100.0** |
| Age (years) |  |  |  |
|  | 5-8 | 9 | 22.0 |
|  | 9-12 | 14 | 34.1 |
|  | 13-17 | 18 | 43.9 |
|  | **Total** | **41** | **100.0** |

Source: SAME/HAN

Regarding the presence of BKPyV infection, most patients were not infected (56.1%). Regarding the type of antiproliferative and calcineurin inhibitor, Mycophenolate Mofetil was used in 37 (90.2%) patients and Tacrolimus in 40 (97.6%) (Table 2).

**Table 2.** Distribution of the sample of children undergoing kidney transplantation in a referral hospital according to the presence of BKPyV infection, type of antiproliferative, calcineurin inhibitor and worsening of creatinine. Salvador - BA, 2009 to 2017.

| **Variables** | **n** | **%** |
| --- | --- | --- |
| **BKPyV infection** |  |  |
| Yes | **18** | **43.9** |
| No | **23** | **56.1** |
| **Total** | **41** | **100.0** |
| **Type of antiproliferative** |  |  |
| Azatioprina | **4** | **9.8** |
| Mycophenolate | **37** | **90.2** |
| **Total** | **41** | **100.0** |
| **Calcineurin inhibitor** |  |  |
| Tacrolimus | **40** | **97.6** |
| Cyclosporine | **1** | **2.4** |
| **Total** | **41** | **100.0** |
| **Worsening of creatinine** |  |  |
| Yes | **10** | **24.4** |
| No | **31** | **75.6** |
| **Total** | **41** | **100.0** |

Source: SAMEHAN
BKPYV: BK Polyomavirus

When comparing data on sex, age and type of antiproliferative agent of patients with the presence or absence of BKPyV infection, no statistically significant difference was identified between these variables as risk factors for Polyoma infection (Table 3).

**Table 3.**
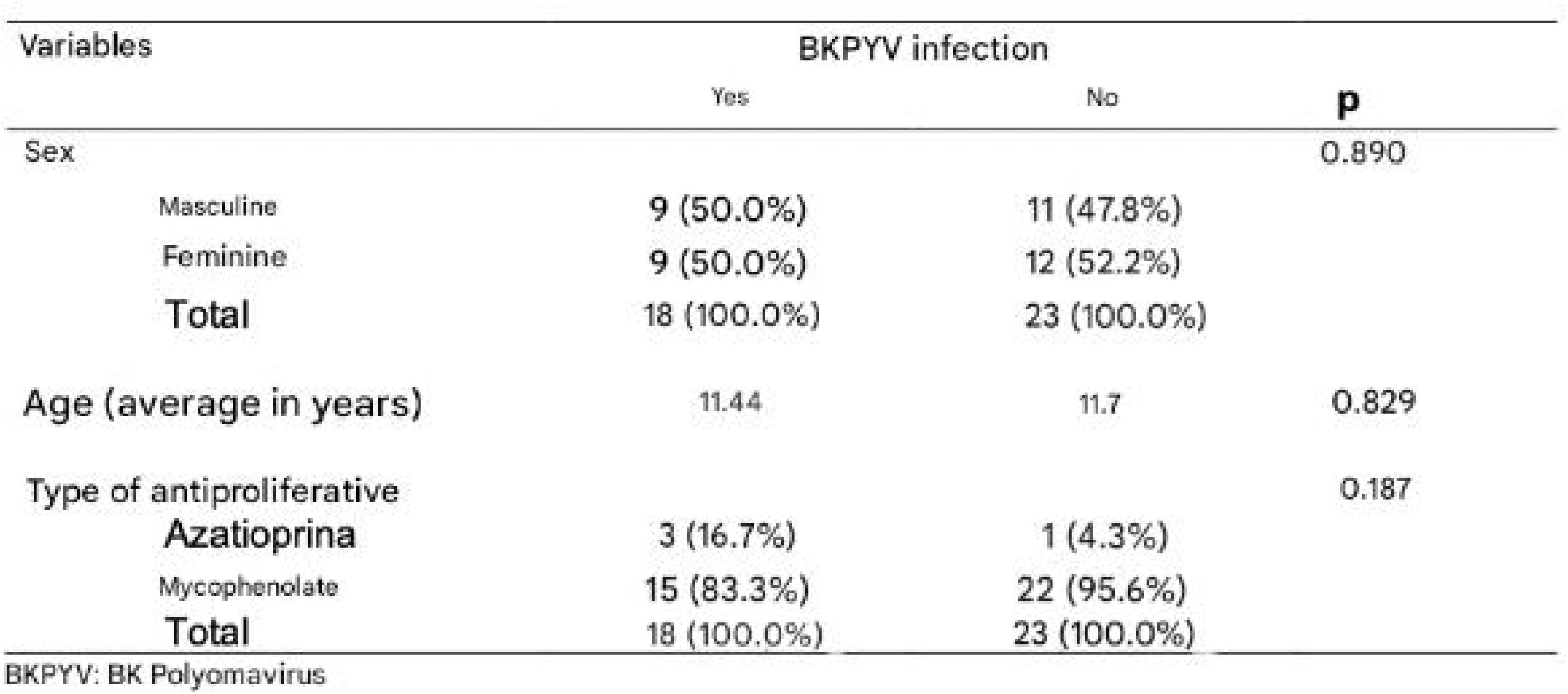
Distribution of the sample of children undergoing kidney transplantation in a reference hospital according to the presence by BKPyV infection. Salvador - BA, 2009 to 2017.

| Variables | BKPyV infection |  | p |
| --- | --- | --- | --- |
|  | Yes | No |  |
| Sex |  |  | 0.890 |
| Masculine | 9 (50.0%) | 11 (47.8%) |  |
| Feminine | 9 (50.0%) | 12 (52.2%) |  |
| **Total** | **18 (100.0%)** | **23 (100.0%)** |  |
| Age (average in years) | 11.44 | 11.7 | 0.829 |
| Type of antiproliferative |  |  | 0.187 |
| Azatioprina | 3 (16.7%) | 1 (4.3%) |  |
| Mycophenolate | 15 (83.3%) | 22 (95.6%) |  |
| **Total** | **18 (100.0%)** | **23 (100.0%)** |  |

BKPyV: BK Polyomavirus

In the analysis of renal function, an increase in creatinine (Appendix B) was observed in infected patients over a period of one year (p = 0.05), thus demonstrating an association between worsening renal function and the presence of infection (Table 4).

**Table 4.** Assessment of renal function of pediatric patients undergoing kidney transplantation in a referral hospital for a period one year and association with the presence or absence of BKPyV infection. Salvador - BA, 2009 to 2017.

| Variables | BKPyV infection |  | p |
| --- | --- | --- | --- |
|  | Yes | No |  |
| Creatinine month 1 (mean in mg/dL) | 0.728 | 0.887 | 0.172 |
| Creatinine month 3 (mean in mg/dL) | 0.800 | 0.896 | 0.368 |
| Creatinine month 12 (mean in mg/dL) | 0.989 | 0.887 | 0.496 |
| Worsening of creatinine |  |  | 0.050 |
| Yes | 7 (38.9%) | 3 (13%) |  |
| No | 11 (61.1%) | 20 (87%) |  |
| **Total** | **18 (100.0%)** | **23 (100.0%)** |  |

BKPyV: BK Polyomavirus

## 6. DISCUSSION

In kidney transplantation, the incidence of BK polyomavirus (BKPyV) infection is high. Given that this infection can result in nephropathy and urethral stricture in kidney recipients, BKPyV is considered a significant cause of premature graft failure after transplantation. 2,13

The present study demonstrated that, among the 41 transplant patients, 43.9% were diagnosed with BK polyomavirus infection through PCR viruria or through renal biopsy. In a retrospective, multicenter, international longitudinal cohort analysis, Höcker B et al 13 reported the presence of BKPyV by viruria in 45.7% of their pediatric patients during the 1st year post-transplant, corroborating the result of this study. It is important to emphasize that a higher incidence of BKPyV replication was observed in pediatric patients, which exceeds the rates in adult kidney transplantation reported by other researchers. The findings obtained in adults, therefore, cannot necessarily be considered for the pediatric population. 14,15

When analyzing the epidemiological profile of patients infected with BK Polyomavirus in this study, there was no statistically significant difference in the frequency of men and women (p = 0.89), corresponding to 22% for both sexes. Values similar to these were found by authors such as Höcker B et al 13 and Brennan D et al 16, which does not characterize the variable sex as a risk factor for infection. Analyses performed by Puliyanda D 17 and Höcker B et al 13 reported that the age of the recipient is associated with the replication of BK Polyomavirus, so that younger children have a higher risk of BKPyV viruria and viremia. This probably occurs due to the fact that a significant proportion of younger children are still immunologically incompetent to this virus.13

The results obtained in this study, however, did not demonstrate a statistically significant difference (p = 0.829) between the mean age of the children and the presence or absence of infection. This difference in relation to the studies cited was mainly due to the small size of the sample analyzed - in which there was no adequate distribution of patients with different age groups - with the highest frequency of children (43.9%) between 13 and 17 years old.

The immunosuppressive therapy used in the transplant patients in this study consisted of a combination of prednisone, antiproliferative and calcineurin inhibitor. Mycophenolate was the antiproliferative of choice in 90.2% of cases, to the detriment of Azathioprine, which was used in only 9.8%. Regarding the calcineurin inhibitor, Tacrolimus was present in the therapy of 97.6% of patients, while Cyclosporine was used in 2.4%.

Regarding these immunosuppressive drugs, no statistically significant difference was identified between these variables as risk factors for Polyoma infection (p = 0.187 for the type of antiproliferative and no comparison was made between calcineurin inhibitor, since only one patient used Cyclosporine). This was probably due to the small sample size of this research, given that larger studies, such as those by Höcker B et al 13 , Miettinen J et al 19, Smith J et al 20 and Hirsch H et al 18,21 stated that Tacrolimus-based therapy, as well as greater intensity of immunosuppression, are both factors associated with a higher risk of BK infection.

Since Tacrolimus is most often administered in combination with other immunosuppressants, these authors analyzed the impact of different regimens on BKPyV infection and, according to these analyses, immunosuppression based on Tacrolimus/Mycophenolate was significantly associated with a two-fold higher risk of BKPyV than the regimen based on Cyclosporine/Mycophenolate.13,18,21

To date, the mainstay of therapy and prophylaxis of BK polyomavirus infection and its possible subsequent nephritis (BKVN) is the reduction of immunosuppressive therapy, since there is currently no established antiviral treatment. Given that late diagnosis of BKVN is generally associated with irreversible decline in graft function and that most patients with viremia will progress to BKVN, regular screening for BKPyV reactivation, mainly during the first two years post-transplantation, with subsequent preventive reduction of immunosuppression, represents the procedure most commonly adopted by transplant centers.2,13,14,18,22

Studies state, however, that there is no uniform strategy for how immunosuppressive therapy should be reduced.2,13 In this study, once diagnosed, patients underwent immunosuppression reduction with a reduction in calcineurin inhibitors and, if there was no improvement, the dose of antiproliferatives was also reduced. Although there is no clear evidence, in vitro analyses support this specific change in therapy, with the first step being the reduction of calcineurin inhibitors due to their effect on T lymphocytes.13,23

Authors, such as Varella R et al 2, recommended that careful monitoring of renal function be performed after immunosuppression reduction, due to the risk of rejection. In the present study, serum creatinine measurements were performed in the 1st, 3rd and 12th month post-transplant in all transplant patients. Although an increase in these values was observed in patients infected with BK, there was no statistically significant difference between the two groups (p = 0.172 in the 1st month; p = 0.368 in the 3rd month and p = 0.496 in the 12th month), a result that can be supported by the analysis of Brennan D et al 16, in which a value of p = 0.15 was obtained for the difference between the mean creatinine levels in the BK-positive and -negative groups in the 1st, 3rd, 6th and 12th months post-transplant.

From the point of view of worsening renal function, the criterion established was an increase in creatinine levels 1.5 times the value prior to the diagnosis of infection, which characterizes, according to the Brazilian Society of Nephrology24, an acute kidney injury. Among the infected patients, 38.9% had worsening creatinine levels, while in the uninfected group, only 13% had worsening. Statistical analysis showed a tendency for creatinine levels to worsen in the population infected with polyomavirus (p = 0.05), which has already been demonstrated in other studies, such as those by Ji et al 25 and Cirocco R et al 26, who stated that there was a significant correlation between the presence of BKPyV and worsening renal function, which was indicated by elevated creatinine levels. The limitations of this study include the fact that a secondary data source was used, which is likely to lead to errors in filling out the medical records. In addition, the small sample size generates a certain imprecision and may lead to type I and type II errors. Furthermore, the collection was carried out in a single reference center, giving rise to selection bias and compromising the external validity of the study.

Thus, it can be concluded that, although numerous retrospective and prospective studies in adults are available, published data on pediatric kidney transplant recipients are limited, highlighting the importance of conducting multicenter studies with a significant sample size in this population.

## 7. CONCLUSION

It was observed that there is an incidence of BK Polyomavirus infection of almost 50% in pediatric patients undergoing kidney transplantation, in the period of one year after the surgical procedure. No differences could be demonstrated between the groups with and without BKPyV infection and established risk factors such as sex, age and immunosuppressive therapy used. Furthermore, worsening of renal function was evidenced among infected patients, when compared to the uninfected group, in this period of one year.

## Data Availability

All data produced in the present study are available upon reasonable request to the authors

---

**APPENDIX A.**
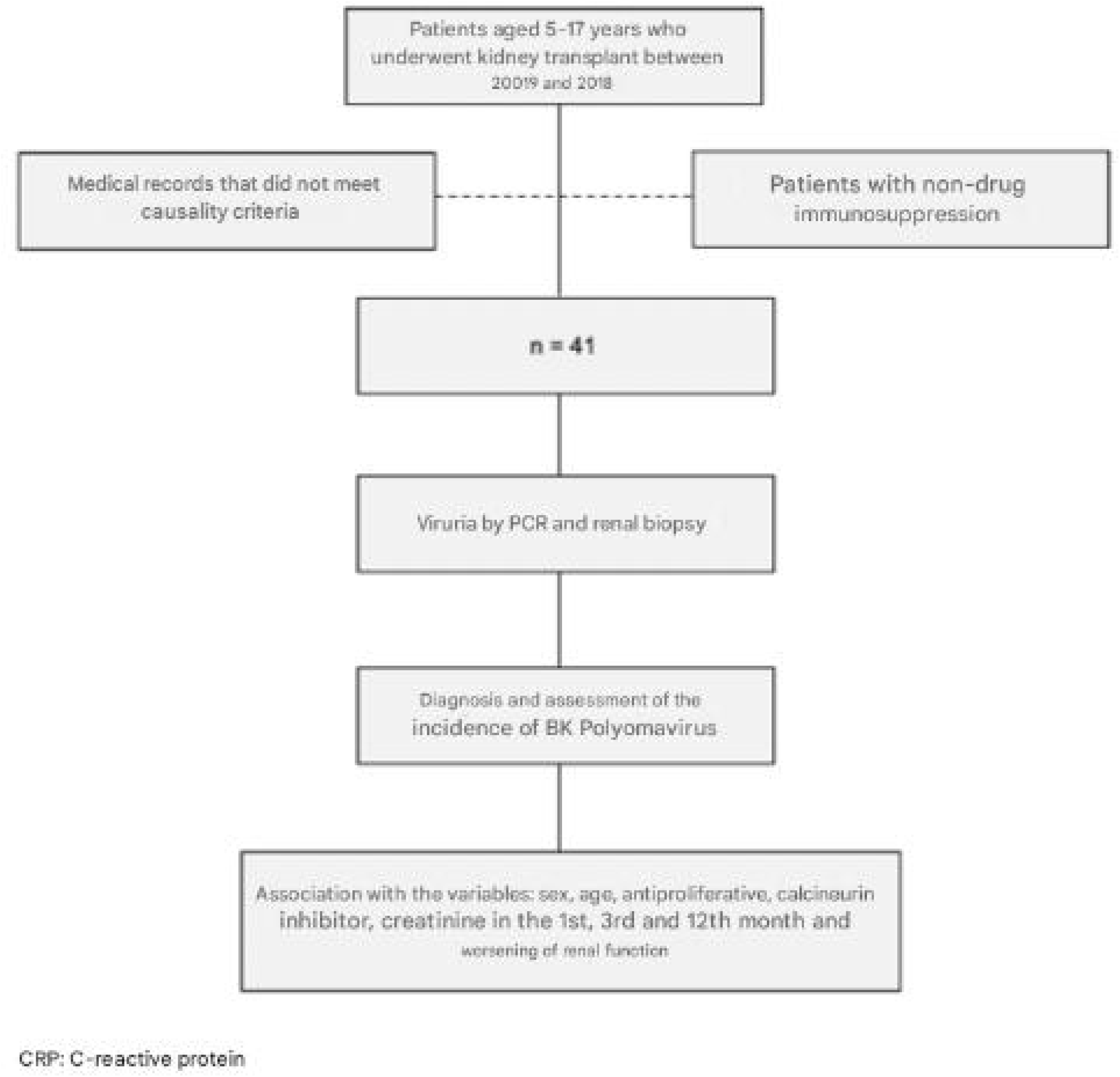
Flowchart of selection and operationalization of collection

**APPENDIX B.**
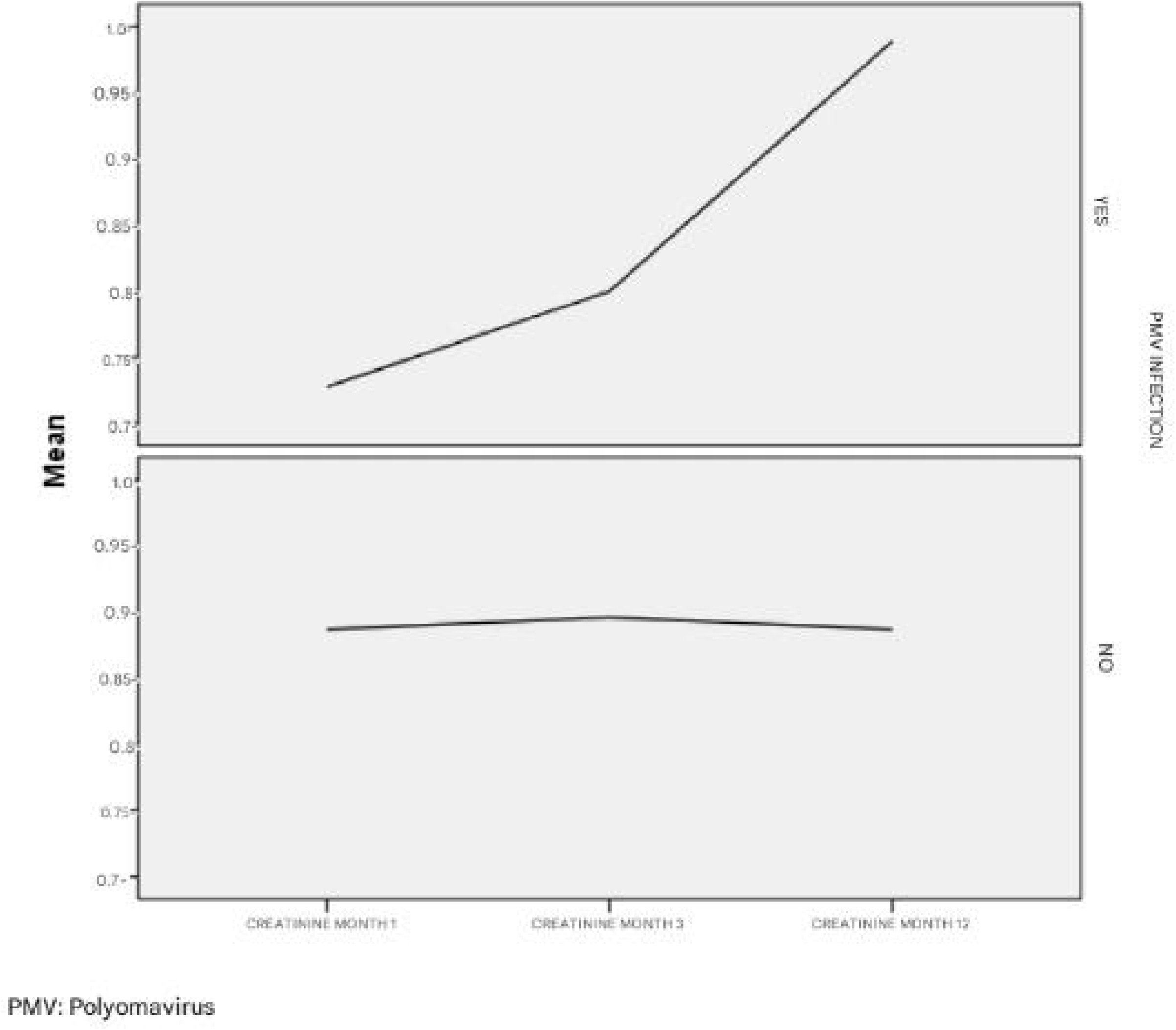
Graph corresponding to the comparison between creatinine values in the 1st, 3^rd^ and 12^th^ month after kidney transplantation, between the infected and non-infected groups.

## Notes

### Competing Interest Statement

The authors have declared no competing interest.

### Funding Statement

This study did not receive any funding

### Author Declarations

Ethics committee/IRB of Hospital Ana Ney gave ethical approval for this work

